# Resource-efficient medical vision language model for dermatology via a synthetic data generation framework

**DOI:** 10.1101/2025.05.17.25327785

**Authors:** Abdurrahim Yilmaz, Furkan Yuceyalcin, Rahmetullah Varol, Ece Gokyayla, Ozan Erdem, Donghee Choi, Ali Anil Demircali, Gulsum Gencoglan, Joram M. Posma, Burak Temelkuran

## Abstract

Vision-language models (VLMs), with their ability to integrate visual and textual information, have enabled unified and interpretable multimodal reasoning. However, developing explainable, image-based artificial intelligence (AI) systems for medicine requires locally deployable models designed to ensure privacy-preserving data workflows. Here, we present SCALEMED (Scalable Clinical Assistants and LEarning for MEDicine), a modular framework that enables the development of locally deployable medical VLMs using small models and synthetic data. The SCALEMED framework integrates clinician data annotation, open-source image-text data collection, synthetic data generation through knowledge transfer using larger VLMs, and fine-tuning of small VLMs to develop domain-specific medical AI systems. As a use case in dermatology, we train a resource-efficient VLM, DermatoLlama, which demonstrates higher success rates in report generation compared to state-of-the-art VLMs across text and image-based evaluation datasets. DermatoLlama, based on Llama 3.2, was trained using DermaSynth, a dataset comprising 1.2 million synthetic text samples generated from 367 expert-crafted seed tasks and 82,379 open-source dermatological images. The SCALEMED framework offers a practical solution for developing explainable and accessible medical AI systems, particularly in resource-constrained healthcare environments.

## Introduction

Large language models (LLMs) are increasingly recognized as transformative tools in healthcare, enabling explainable analysis, structured diagnoses, and personalized treatment plans [1–4]. They have also demonstrated promise in providing multi-modal solutions, such as interpreting radiological images, augmenting clinical decision-making, and streamlining workflows [5–7]. Despite these advancements, widespread adoption of LLMs in medicine still faces significant barriers. These include limited availability of high-quality and diverse datasets [8], stringent privacy regulations [9, 10], and the high computational and workforce costs [11, 12] required for training and deployment of LLMs in clinical settings. Such constraints cause significant challenges for the adaptation of LLMs to real-world and resource-limited environments. Therefore, developing resource-efficient and privacy-preserving solutions allowing scalability to multiple fields is essential for effectively integrating LLMs into healthcare practice.

Building task-focused artificial intelligence (AI) assistant systems (‘agents’) and generating synthetic data, with or without ground-truth, have emerged as powerful strategies for overcoming the high computational and extensive data demands of LLMs in healthcare [13–15]. By transferring knowledge from a large teacher model to a lighter, more efficient student model, ‘knowledge transfer’ reduces the resources needed for training and inference. Collaborative AI assistants (‘multi-agents’) distribute complex tasks among specialized assistants that communicate and cooperate, enabling medical AI to process diverse inputs (e.g., images and texts) while maintaining flexibility and scalability [16–19]. Despite these promising methodologies, current evaluation practices and data-handling approaches rarely harness LLMs to their full potential. The field lacks a unified framework that incorporates knowledge transfer, multi-agent designs, and synthetic data generation to address these real-world constraints [20].

Moreover, privacy concerns and data scarcity remain pressing issues. Many organizations are not allowed to use AI models hosted on external servers due to constraints for sharing confidential medical data. In addition, researchers require access to high-quality processed data to build sufficiently representative datasets that capture the full spectrum of patient variation. A solution addressing these issues could accelerate the deployment of LLM-driven healthcare tools, enabling robust performance across medical specialties while safeguarding patient confidentiality and optimizing resources for institutional usage.

To address these challenges in developing robust, privacy-preserving, and scalable AI systems for healthcare, we propose a data-centric framework, SCALEMED (Scalable Clinical Assistants and LEarning for MEDicine) (Fig. 1). Unlike traditional approaches that rely on tightly controlled datasets and static evaluation formats, SCALEMED integrates synthetic data generation (Fig. 1a), leveraging open-access (OA) databases and PubMed (PM) literature to create a diverse, representative dataset of medical information, with a specific use case in dermatology. This dataset is enriched through flexible instruction methods, ranging from AI-generated clinical questions derived from high-quality references (self-instruct) to medical knowledge-based prompts and anamnesis-style narratives. These flexible instruction methods allow additional ones to be integrated and expose vision enabled LLMs (‘vision language model (VLM)’) to a wide array of clinical scenarios. SCALEMED also highlights critical licensing considerations, guiding researchers toward safe and compliant utilization of publicly available data. SCALEMED allows users to generate synthetic evaluation data by using state-of-the-art (SOTA) LLMs, which can be edited using our open-source annotation tool, AnnotatorMed. To enhance computational efficiency, SCALEMED can be integrated with techniques such as Low-Rank Adaptation (LoRA) and Quantized LoRA (QLoRA) during model fine-tuning. These techniques enable efficient adaptation of OA models, such as Llama and its vision variants, significantly reducing the computational load without sacrificing model performance. SCALEMED further refines these models through knowledge transfer, while protecting patient privacy by enabling local data annotation and model development on lowcost computers. An iterative, modular design ensures the framework is adaptable, explainable, and scalable, facilitating multi-agent systems in which specialized agents can collaborate.

**Figure 1:**
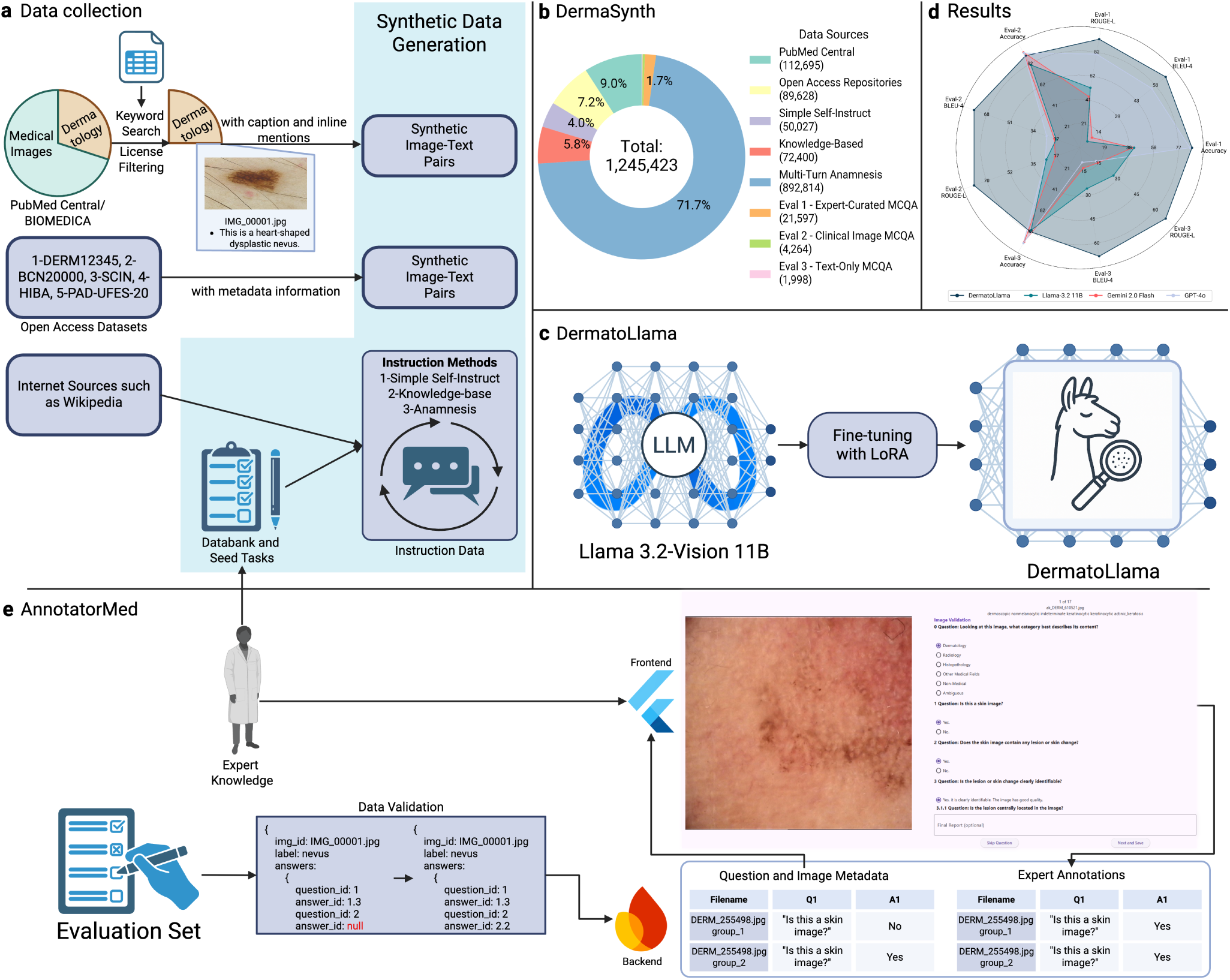
Overview of SCALEMED framework. **a,** Data collection pipeline of DermaSynth—1.2 million data samples based on PubMed, open access datasets and instruction-follow methods. **b,** DermaSynth data size and its data sources. **c,** DermatoLlama was fine-tuned on Llama-3.2-11B-Vision using LoRA optimization method. **d,** DermatoLlama can reach GPT-4o, Gemini 2.0 Flash in accuracy, and can surpass all models in report generation for all evaluation sets. **e,** AnnotatorMed is a free and open source tool for data annotation in clinical context.

We demonstrated the application of the SCALEMED framework in dermatology, a domain currently lacking sufficiently diverse, high-quality datasets essential for advanced clinical reasoning [21, 22]. Using SCALEMED, we created DermaSynth, a synthetic dataset tailored specifically for dermatology, comprising approximately 1.2 million text samples (Fig. 1b). DermaSynth includes synthetic samples generated using 56,258 images sourced from PubMed articles and 44,814 images from open access datasets. We enriched this synthetic data through clinical instruction-following (IF) methods where LLM generates new samples based on reference samples (‘seed tasks’) (Methods). Our seed tasks facilitate effective knowledge transfer from domain sources to our model. These methods help to replicate real-world dermatological challenges at scale, enabling the development of our VLM, DermatoLlama, to achieve deep clinical insight with comprehensive report generation. A code-driven evaluation pipeline generates synthetic question-answer (QA) pairs, including image-based queries, allowing systematic performance assessment without relying on sensitive patient data. AnnotatorMed, an open-source annotation tool developed within this framework, preserves privacy by supporting local development on standard hardware, while its modular, iterative, and hierarchical design enables cost-efficient deployments (Fig. 1e). By consolidating synthetic data generation, model fine-tuning, open-source tools, expert annotation, and flexible deployment, SCALEMED aims to deliver a transparent, ethical, and high-performing ecosystem for real-world clinical applications. Our findings show SCALEMED’s potential as a scalable, privacy-preserving, domain-specific, and cost-effective framework capable of significantly advancing medical AI models.

## Results

### SCALEMED Framework Implementation and DermaSynth

We implemented SCALEMED framework (Fig. 1) to generate a large-scale, multifaceted synthetic dataset for dermatology, DermaSynth, using three self-instruct methods based on OA image datasets (Fig. 2a), PM articles (Fig. 2b), and IF dataset (Fig. 2c). DermaSynth comprises approximately 1.2 million synthetic samples (Fig. 1b), integrating visual question-answer (VQA) pairs derived from metadata-rich OA datasets on two common dermatological image modalities (n = 26,888 QA pairs of 13,444 clinical images and n = 62,740 QA pairs of 31,370 dermoscopic images) and the clinically relevant BIOMEDICA dataset (n = 112,695 QA pairs of n = 37,565 images, no modality information) [23] based on PM articles (Methods). To construct the synthetic IF dataset, we initially designed seed tasks for guidance covering diverse clinical scenarios. Synthetic data was generated through self-instruction method categorized into: (1) simple IF QA (n = 190 seed tasks (Supplementary Data 1), n = 50,027 synthetic samples), (2) knowledge-based QA (n = 40 seed tasks (Supplementary Data 2), n = 72,400 synthetic samples), and (3) anamnesis-style case studies (n = 137 seed tasks (Supplementary Data 3-5), n = 892,814 text samples of n = 9,963 synthetic cases). Additionally, we created three evaluation sets (DermaBench) to assess VLMs: dermoscopic image analysis in multi-turn conversations (Eval1, n = 21,597) (Fig. 1e and Methods), clinical image analysis with report generation based on PubMed images (Eval2, n = 4,264), and general dermatology knowledge (Eval3, n = 1,998). This data-centric approach established comprehensive baseline datasets for training and evaluating a specialized dermatology VLM, DermatoLlama (Supplementary Movie 1).

**Figure 2:**
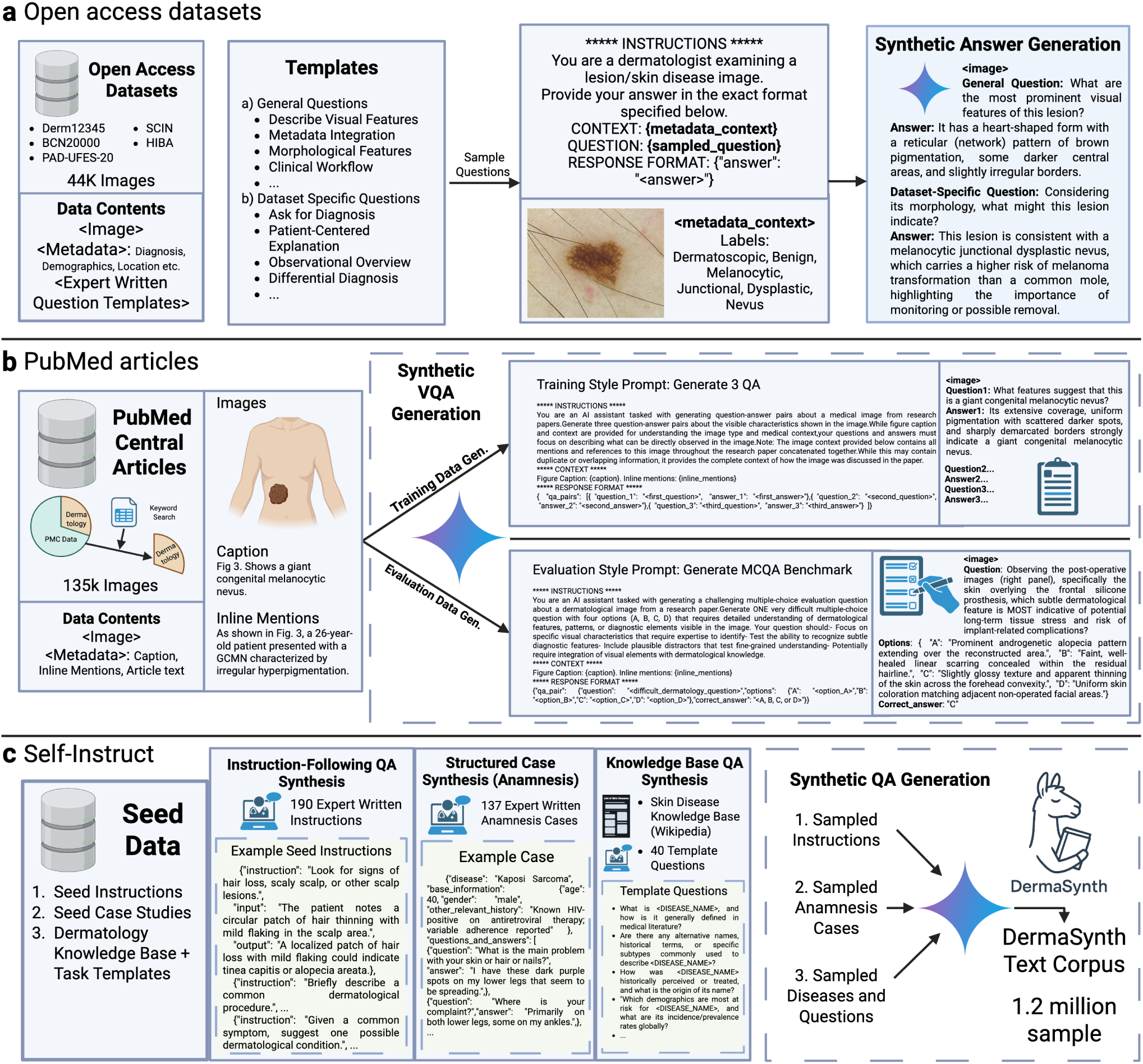
Overview of synthetic data generation workflow. **a,** 44k open access dermatology images with metadata were used to generate grounded QA pairs (medium-trust/hallucination). **b,** 37k open access PubMed images with captions and inline mentions were used to generate descriptive QA for training and MCQs for evaluation (high-trust, low-hallucination). **c,** 1,014k Synthetic instruction-following cases were generated based on self-instruct, knowledge base and anamnesis to cover clinical reasoning (low trust, high-hallucination).

### DermatoLlama Performance on Benchmark Evaluations (DermaBench)

We evaluated our fine-tuned model, DermatoLlama -Llama-3.2-11B-Vision fine-tuned on DermaSynth-against the base Llama-3.2-11B-Vision and SOTA models (GPT-4o, knowledge cutoff: October 2023; Gemini 2.0 Flash, knowledge cutoff: August 2024). The comparison was conducted across three distinct benchmarks, DermaBench (Eval1, Eval2, Eval3), each targeting different aspects of dermatological reasoning and knowledge. Additionally, we performed targeted evaluations to assess the quality and effectiveness of each distinct synthetic data generation pipeline within DermaSynth (Fig. 2). For this analysis, we trained separate model instances using progressively adding more data: 10k (OA), 50k (OA), 100k (OA), 200k (OA+PM), all samples (OA+PM+IF). We then evaluated each trained instance on evaluation benchmarks (Eval1, Eval2, Eval3). This targeted approach validated the contribution of each data generation strategy. Detailed results comparing the DermatoLlama models trained on various dataset sizes with the SOTA models are presented in (Table 1) and (Fig. 3).

**Figure 3:**
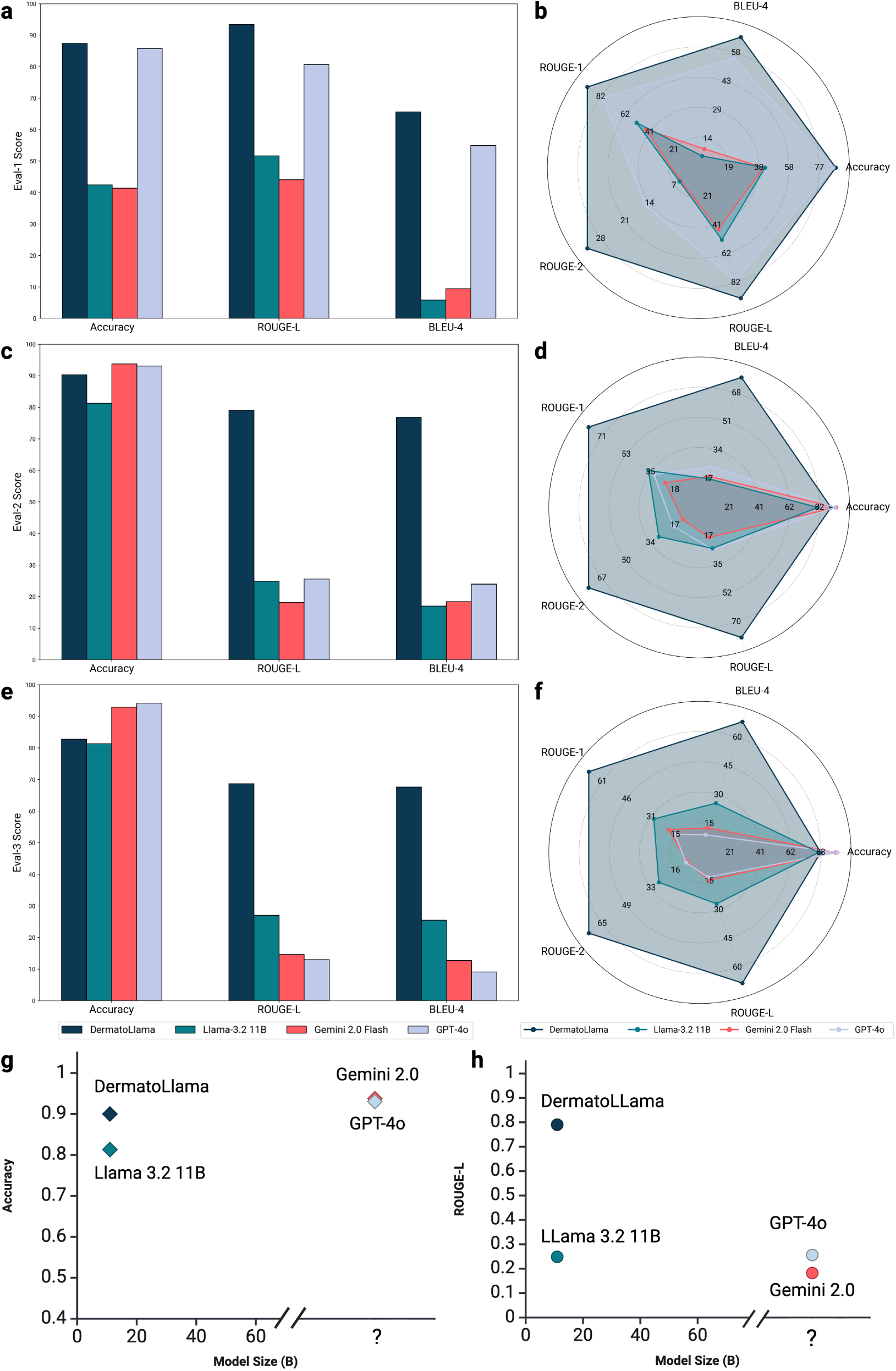
Comparison of results. **a,b,** Eval1 - classification and report generation performance of DermatoLlama. **c,d,** Eval2 - classification and report generation performance of DermatoLlama. **e,f,** Eval3 - classification and report generation performance of DermatoLlama. **g,h,** Eval2 - Scatter plots of accuracy and ROUGE-L versus parameter count showcase DermatoLlama’s - small-but-mighty-profile—delivering large-model performance from a compact footprint.

**Table 1:**
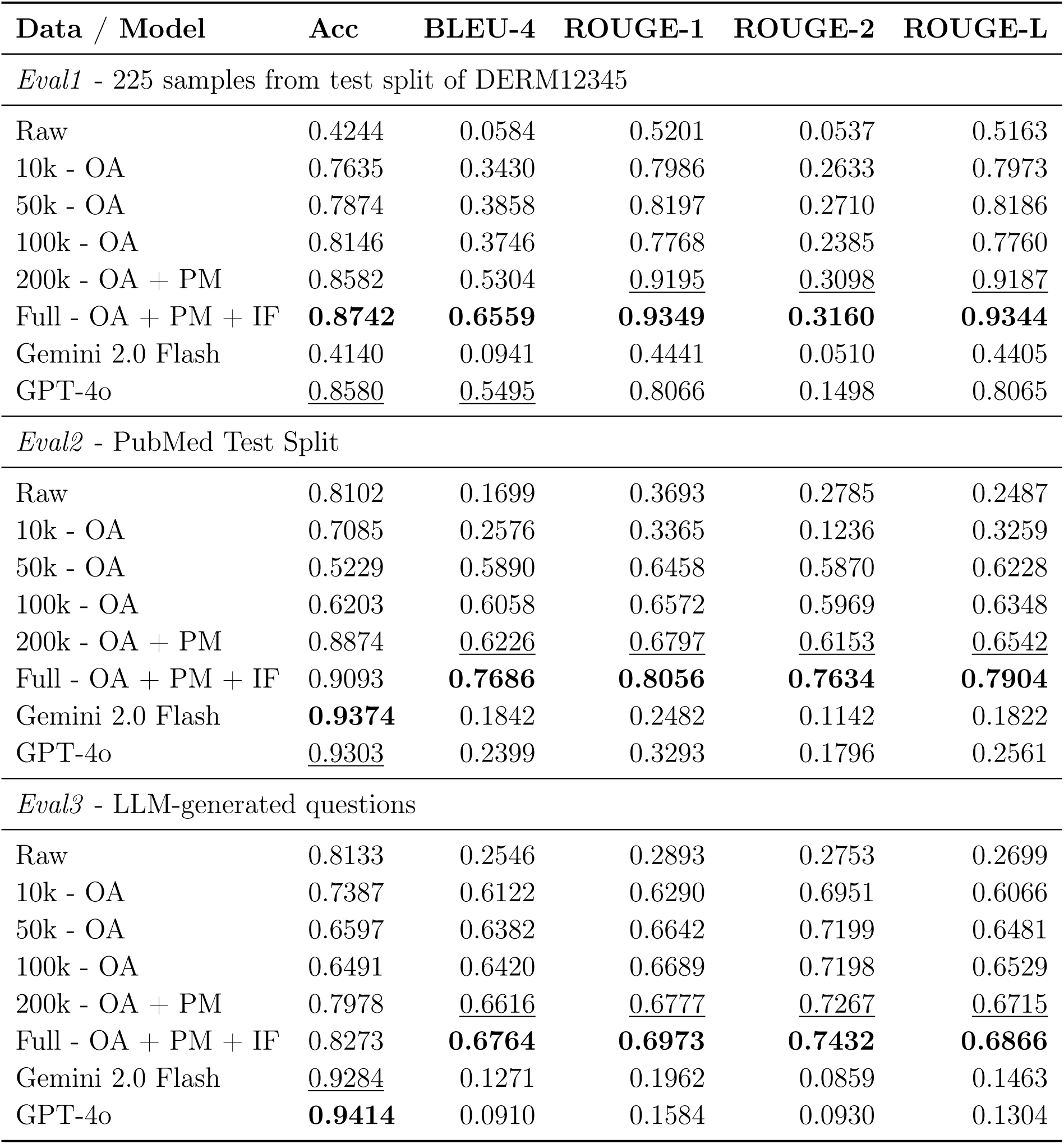
Classification and report generation performance of DermatoLlama with different variants and SOTA models, GPT-4o and Gemini 2.0 Flash. OA: Open access images with QA pairs, PM: Open access PubMed images with QA pairs, IF: Text-based instruction-following samples. The highest metric across all models per evaluation set is given in **bold**, and second highest is <u>underlined</u>.

#### Evaluation on Eval1: Multi-Turn Dermoscopic Image Analysis

We designed a hierarchical question set for the two-step pattern analysis [24] to evaluate the systematic diagnosis capability of AI models (Table 1) and Methods). This question set comprises 93 root questions and 320 total questions (Supplementary Data 6-8) organized into four clinical-dermoscopic subsections: basic clinical context, clinical context, dermoscopic context, and final diagnosis (Supplementary Table 1). Eval1 contains 225 expert-annotated images from the test split of DERM12345, featuring single-choice, multi-choice, and open-ended items requiring chained clinical reasoning. The DermatoLlama - Full model (trained on all samples of DermaSynth) demonstrated significantly improved performance compared to the base Llama model. DermatoLlama achieved an accuracy (Acc) of 0.87, along with text-generation scores of BLEU-4 = 0.66, ROUGE-1 = 0.94, ROUGE-2 = 0.32, and ROUGE-L = 0.93. In contrast, the base Llama model had considerably lower scores (Acc = 0.42, BLEU-4 = 0.06, ROUGE-1 = 0.52, ROUGE-2 = 0.05, ROUGE-L = 0.52). DermatoLlama also outperformed GPT-4o (Acc = 0.86, BLEU-4 = 0.55, ROUGE-1 = 0.81, ROUGE-2 = 0.15, ROUGE-L = 0.81) with better accuracy and report generation, and significantly surpassed Gemini 2.0 Flash (Acc = 0.41, BLEU-4 = 0.09, ROUGE-1 = 0.44, ROUGE-2 = 0.05, ROUGE-L = 0.44) across all metrics. Example reports of ground truth, DermatoLlama, and the base Llama model are presented in (Fig. 4). Other DermatoLlama models, trained on different sample sets of DermaSynth (OA, OA+PM, OA+PM+IF), also consistently outperformed the base Llama model, confirming that converting clinical data into structured descriptions provides high-fidelity supervision. The additional data obtained from PubMed and IF approaches demonstrate complementary benefits from combining diverse data sources.

**Figure 4:**
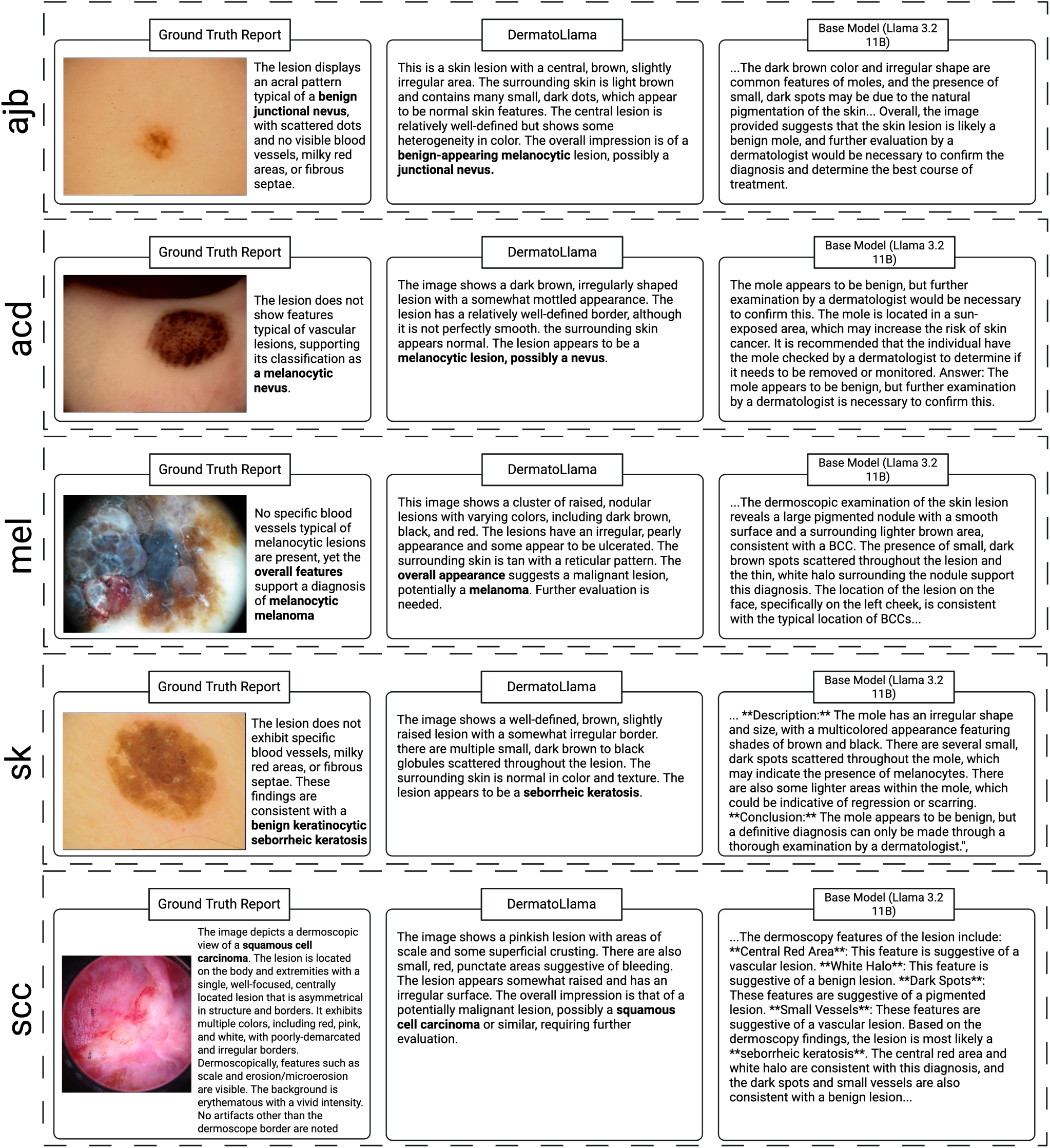
Example reporting results. Dermoscopic cases—ajb (acral junctional banal nevus), acd (acral compound dysplastic nevus), mel (melanoma), sk (seborrheic keratosis), scc (squamous cell carcinoma)—with the ground-truth report (left), DermatoLlama output (centre), and base Llama output (right). DermatoLlama descriptions align more closely with the reference than the base Llama model.

#### Evaluation on Clinical Multiple-Choice VQA (Eval2)

The Eval2 benchmark tested the models’ ability to answer multiple-choice question answers (MCQA) based primarily on clinical images and associated text from the BIOMEDICA dataset (Table 1) [23]. DermatoLlama - Full achieved an accuracy of 0.91, along BLEU-4 = 0.77 and ROUGE-L = 0.79. This model surpassed the base Llama model (Acc = 0.81, BLEU-4 = 0.17, ROUGE-L = 0.25). Although GPT-4o (Acc = 0.93) and Gemini 2.0 Flash (Acc = 0.94) demonstrated slightly higher classification accuracy, DermatoLlama outperformed both in BLEU-4 and ROUGE-L scores, indicating superior report quality. Additionally, the performance of DermatoLlama - OA (Acc = 0.62, BLEU-4 = 0.61, ROUGE-L = 0.60) and DermatoLlama - OA+PM (Acc = 0.89, BLEU-4 = 0.62, ROUGE-L = 0.65) confirms the value of synthetic QA derived from PM sources. These results highlight that broader data exposure, as utilized in the full model, leads to improvements in both accuracy and text scores. These findings demonstrate that synthetic data derived from high-trust PM sources significantly improves the model’s ability to interpret dermatological figures in the clinical literature. In addition, increased data diversity further enhances both diagnostic accuracy and language quality.

#### Evaluation on Text-Only Dermatology Knowledge MCQs (Eval3)

To assess the transfer of textual dermatological knowledge without images, we used the Eval3 benchmark, comprising text-only MCQA (Table 1). DermatoLlama - Full achieved an accuracy of 0.83, BLEU-4 = 0.68, and ROUGE-L = 0.69, surpassing the base Llama model (Acc = 0.81, BLEU-4 = 0.26, ROUGE-L = 0.27) in both accuracy and report generation metrics. While GPT-4o (Acc = 0.94) and Gemini 2.0 Flash (Acc = 0.93) performed better in classification accuracy, their report generation quality remained low (BLEU-4 ≤ 0.12; ROUGE-L ≤ 0.15), highlighting DermatoLlama’s superior explanation capabilities beneficial for clinicians. DermatoLlama’s significant improvement over self-instruction strategies successfully created a diverse and accurate dermatological knowledge base, effectively embedding domain-specific textual understanding into the language component of the VLM.

### Resources Released

We make the following assets publicly available:

- **DermaSynth** - a dataset of approximately **multimodal 1,2 million** synthetic data points around 250k image-text pairs (clinical + dermoscopic) in both Hugging Face Datasets and JSON formats (Fig. 1a).
- **DermatoLlama 1.0** - an 11B parameter, long-context (2,048 tokens) vision-language model fine-tuned with LoRA adapters on a DermaSynth mixed real + synthetic dermatology set to process longer clinical texts and images (Fig. c).
- **AnnotatorMed** - an open-source, locally deployable, and user-friendly web and mobile application. AnnotatorMed is a Flutter application with a Firebase backend for hierarchical data annotation, clinician test, and optional reinforcement learning from human feedback (RLHF) [6, 25] logging, an approach to improve AI performance with relatively small feedbacks (Fig. 1e).
- **SCALEMED Framework** - end-to-end utilities (data scraping, synthetic generation, fine-tuning, evaluation) and a Colab notebook illustrating how to adapt the pipeline to any medical specialty (Fig. 1).

All source codes are hosted on github.com/DermaVLM. The DermatoLlama model (trained on the full DermaSynth dataset) along with its checkpoints, training, and evaluation datasets, are available on Hugging Face (huggingface.co/DermaVLM). Additionally, the DermatoLlama model is directly accessible via the Hugging Face Space named DermatoLlama. Use of the dataset, model, and code is restricted to **non-commercial research**, in accordance with the licensing terms documented in the repository.

## Discussion

Deploying LLMs in resource-constrained organizations remains challenging due to limited data availability, stringent privacy regulations, and high costs of development and maintenance [9, 26–30]. We developed SCALEMED, a resource-efficient and privacy-preserving framework that can train and deploy successful AI models, addressing these challenges faced in medicine. SCALEMED is an integrated framework specifically designed as a modular, data-centric workflow for developing medical VLMs. SCALEMED integrates synthetic data generation, an open-source data annotation tool, and large-scale fine-tuning into a unified, scalable, end-to-end pipeline (Fig. 1).

We demonstrated that: (i) using synthetic data enriched with targeted clinical instructions significantly improves both classification and report generation performance of our model, DermatoLlama; (ii) each synthetic data source (OA images, PM-derived data, and IF methods) contributes distinct and additive performance gains; and (iii) the full variant of DermatoLlama matched or surpassed commercial, closed-source SOTA models (GPT-4o and Gemini 2.0 Flash) in report generation metrics and achieved comparable classification accuracy. DermatoLlama surpasses the SOTA models on the evaluation set, Eval1 which is based on DERM12345, published after the release of SOTA models used in this study. On Eval2, containing images potentially present in the SOTA models’ training datasets, DermatoLlama has also superior report generation metrics and attains comparable accuracy. Furthermore, when assessed on questions generated by Gemini 2.0 Flash (Eval3), DermatoLlama still reaches an accuracy of 83%. Across all three evaluation sets, DermatoLlama achieved the highest scores in every report generation metrics, consistently outperforming GPT-4o, Gemini 2.0 Flash, and the base Llama model (Fig. 3).

DermatoLlama fine-tuned with smaller sample sizes (10k-50k) did not converge to optimal performance, generating lengthy and convoluted outputs that inflated the amount of produced text. DermatoLlama fine-tuned with all samples (1.2 million) learned to generate clinically relevant responses, eliminating unnecessary text, and significantly improving report quality. Compared to the base Llama model, DermatoLlama’s BLEU-4 score is higher by 11.2× on Eval1, 4.5× on Eval2, and 2.65× on Eval3 (Table 1). Compared to Gemini, BLEU-4 is higher by 6.97× on Eval 1, 4.17× on Eval 2; and 5.32× on Eval3 (Table 1). DermatoLlama’s improved conciseness significantly enhances computational efficiency during both training and inference, providing a critical advantage for daily clinical use. Unlike commercial models, which often generate overly detailed, free-form responses resulting in lower text-quality metrics, DermatoLlama’s targeted fine-tuning yields concise and domain-specific answers (Supplementary Figs. 4-8 and Supplementary Data 9-10). This leads to superior report generation metrics and better alignment with clinical requirements, ultimately improving usability and reliability in medical practice.

In evaluations, we observed consistent performance improvements as we incrementally introduced larger and more diverse datasets during fine-tuning. We increased the data size from thousands of open-access image-text pairs to hundreds of thousands of enriched image-text pairs incorporating PM contexts and IF samples. Starting from an untrained baseline, each increase in data volume yielded higher evaluation metrics. The 50k, 100k, and 200k configurations each demonstrated notable improvements over earlier training stages, while the DermatoLlama-Full achieved the highest overall scores. DermatoLlama’s performance plateaued around 200k samples; however, supplementing with synthetic data significantly expanded its effective knowledge base and enhanced its overall performance. These findings highlight the value of integrating multiple data sources, particularly enriched with synthetic data. IF samples, introduced at the million-sample scale, substantially improved the model’s ability to handle open-ended queries, thereby refining both diagnostic accuracy and report quality.

A critical insight from our dermatology use case is the importance of high-quality synthetic datasets. In settings where real-world data is scarce or cannot be easily shared due to privacy concerns, synthetic data can fill gaps by simulating a broad range of clinical presentations. Our experiments reinforced that the diversity and representativeness of synthetic samples are essential. However, synthetic data should be complemented by clinically grounded resources, such as PM articles. Additionally, we found that seed tasks and databanks can be rapidly created with minimal yet strategic clinician input, streamlining the generation of high-quality synthetic data. This approach reduces the overall burden on domain experts while ensuring clinically relevant outputs.

Recent research in medical AI has focused on developing generic foundation models [4, 31, 32] such as MONET [33], PANDERM [34], DERM1M [35], as well as pretrained models such as SkinGPT-4 [36]. These models primarily emphasize data annotation and short-context diagnostic tasks, falling short of addressing critical clinical needs such as comprehensive report generation. From a clinical perspective, there is an increasing demand for decision support systems that can handle longer contexts, including comprehensive patient histories and detailed narratives [37, 38]. DermatoLlama, a fine-tuned 11-billion-parameter VLM, addresses this gap by generating longer sequences and detailed clinical reports. Moreover, due to its smaller size and reduced computational demands through LoRA approach, DermatoLlama can be efficiently deployed on a single, low cost Nvidia RTX4090 GPU, requiring an initial setup cost of approximately $2,000. The combined cost of synthetic data generation, model fine-tuning, and comprehensive evaluation for this work was under $500. This demonstrates that high-quality, domain-specific models can be developed cost-effectively, making them accessible to institutions with limited budgets, such as academic and clinical settings.

To address the challenges of computational cost, privacy concerns, and scalability, we propose utilizing LoRA-based adapters to efficiently fine-tune LLMs for specialized medical models. Multiple condition-specific adapters fine-tuned with LoRA (e.g., for rare dermatological diseases) can be swapped in and out without duplicating the entire model. This model can also be deployed in cloud environments equipped with GPUs, such as Google Colab with Nvidia A100 GPUs. A similar framework, LLaVa-Med [27], used 8×A100 GPUs for training, incurring an estimated cost of $400-600 for training alone, in addition to significant workforce. This cost saving could be quite significant considering the cumulative cost of multi-agent systems needed in this training for each department and hospital. We integrated LoRA adapters to efficiently fine-tune our model for specialized medical functions while preserving high-quality report generation and complex instruction-following abilities. Our framework also supports privacy-preserving local refinement using proprietary data, allowing institutions full control over data access. By leveraging synthetic data and knowledge transfer, we demonstrated that clinically accessible-size models can achieve clinically relevant performance without the need for massive architectures, pre-training and supervised fine-tuning (SFT). Beyond diagnostic and treatment tasks, this model’s high-quality text generation capabilities enable it to handle additional tasks, such as simplifying medical explanations for laypersons and providing supportive responses to stressed patients. This capability of model underscores its broader applicability beyond clinical use cases and presents a potential to reduce clinicians’ workload by improving the generation and interpretability of clinical documentation.

We demonstrate with AnnotatorMed how a purpose-built, free, open-source, and locally deployable tool can remove one of the largest bottlenecks in medical-AI development: secure, clinician-driven data curation. AnnotatorMed can run on local infrastructure (e.g., hospital servers) behind a firewall, ensuring sensitive images and reports remain securely within institutional storage. This setup enhances security and still enables collaborative labelling projects. AnnotatorMed incorporates a hierarchical workflow specifically designed for cases where a single clinical finding can branch into multiple differential diagnostic pathways (Supplementary Figs. 1 and 2). This hierarchical workflow is particularly effective in our use-case of dermatology, reducing screen clutter and increasing annotation speed. For instance, in Eval1, AnnotatorMed helped clinicians decrease the number of required annotations from 72,000 to 21,597 (Fig. 1e). Beyond labelling, AnnotatorMed can be used to capture RLHF annotations [6] or serve as a clinician test platform, allowing experts to rank, correct, or critique model outputs (Supplementary Fig. 3). Furthermore, the dermatology-specific hierarchical workflow can be adapted to other specialties with some domain-specific adjustments. By unifying multimodal input, flexible question logic, and secure on-premises deployment, AnnotatorMed provides a reusable architecture for high-quality data generation in clinical AI workflows.

There are certain limitations in our study that advancements in the rapidly evolving field of AI may help address. Although synthetic data can significantly increase dataset size, it may fail to capture subtle features, particularly those associated with rare or advanced cases [39, 40]. Additionally, despite the low resource cost of training and deployment, clinical experts are still required to validate and annotate large volumes of generated data, especially for diagnostic models. While our trained model contains 11 billion parameters, making it less resource-intensive than trillion-parameter alternatives, it may still lack the advanced reasoning capabilities required for more complex clinical tasks. However, this reasoning capacity is destined to improve due to ongoing improvements in optimization methods, knowledge transfer, and incremental training. Furthermore, we emphasize that extensive safeguards, such as external clinical validation, bias audits across diverse populations, adversarial stress testing, and robust privacy controls, have not yet been implemented. These measures are crucial for safe clinical deployment, which demands formal regulatory review and rigorous prospective evaluation. Consequently, the current model remains a research prototype and is not yet suitable for guiding individual patient care.

Our framework can seamlessly adapt to evolving clinical needs and diverse medical domains by providing a strong basis for developing advanced AI systems. It supports the creation of multi-agent systems capable of distributing specialized tasks -such as image interpretation, textual reasoning, or patient triage-among smaller, dedicated sub-models [41].

Integrating SCALEMED with methods such as RLHF [6] and retrieval-augmented generation (RAG, a technique enabling AI systems to retrieve reliable medical information) [42] allows the development of AI systems that can mitigate AI hallucinations and improve contextual awareness [43]. These methods can be used to develop resource-efficient models by extending their knowledge bases without significantly increasing parameter counts. In addition, establishing domain-ready baseline models empowers individual institutions to tailor AI solutions to their specific patient populations. Combining SCALEMED with complementary frameworks such as MONET [33], along with its automated concept annotation capability, can improve interpretability and clinical success. The hierarchical approaches used in Eval1 can similarly enhance AI assistant performance in medical applications by guiding researchers toward more effective strategies. An example would be a ‘hierarchy-of-thought’ method, which we hypothetically name in analogy to existing ‘chain-of-thought’ methods used in LLMs [44].

In summary, we introduce a locally deployable framework, SCALEMED, featuring a single medical assistant for dermatology (DermatoLlama), a synthetic dataset (DermaSynth), and a data annotation tool (AnnotatorMed). Our framework overcomes the complexity of adapting trillion-parameter LLMs, enabling clinically applicable solutions compatible with low-cost hardware. This approach, reaching current SOTA model performance, can accelerate the adoption of AI in healthcare by providing a scalable, privacy-preserving, and efficient framework. SCALEMED effectively addresses both current clinical tasks and evolving future needs, accommodating the increasing complexity of healthcare demands.

## Methods

### Data

#### Overall Description

In the initial phase of our framework, SCALEMED focuses on data gathering, task structuring, and synthetic dataset creation. After merging three individual pipelines, we assembled a comprehensive dataset consisting of 112,695 image-text pairs from PubMed (high-trust data source), 89,628 image-text pairs from open-access repositories providing limited metadata (as a medium-trust data source), and synthetic instruction following data (low-trust data source). The synthetic instruction following data comprised 50,027 self-instruct prompts derived from 190 seed tasks (Supplementary Data 1), 72,400 knowledge-based tasks covering approximately 2,700 diseases generated from 40 seed tasks (Supplementary Data 2), and 9,963 multi-turn anamnesis interactions created from 137 disease-specific seed tasks based on 104 fixed questions (Supplementary Data 3-5). This integrated approach streamlines the workflow, enabling researchers to progress seamlessly from raw data sources to a carefully curated synthetic dataset, which serves as the foundation for subsequent model training and evaluation. Details of the data generation process are shown in (Fig. 2). No human participants or personally identifiable data were involved in this study. All image-text pairs, prompts, and derived synthetic data originate from publicly available open-access repositories and literature, thereby obviating the need for additional ethical approval. All licenses (exclusively commercially available sources) and usage rights were verified to ensure no patient privacy or confidential information was compromised. The dataset specifications are detailed in (Table 2).

**Table 2:** Characteristics of DermaSynth dataset.

| Data Source / Task | # Samples | Notes |
| --- | --- | --- |
| PubMed image-text pairs | 112,695 | 37,565 images with three QA pairs |
| Open Access Repositories | 89,628 | 44,814 images with two QA pairs |
| DERM12345 (Train) | 9,860 | Dermoscopic |
| DERM12345 (Test) | 2,485 | Dermoscopic |
| BCN20000 (Train) | 11,761 | Dermoscopic |
| BCN20000 (Test) | 6,075 | Dermoscopic |
| PAD-UFES-20 | 2,292 | Clinical |
| SCIN | 10,858 | Clinical |
| HIBA | 1,572 | 1,270 Clinical, 302 Dermoscopic |
| *Subtotal* | 44,814 | 13,444 Clinical, 31,370 Dermoscopic |
| Self-Instruct | 50,027 | 190 seed task |
| Knowledge-based tasks | 72,400 | 40 seed task, based on Wikipedia [49] |
| Multi-turn anamnesis | 892,814 | 9,963 multi-turn case and 137 seed task |
| DermaBench |  |  |
| Eval1 | 21,597 | 225 samples of DERM12345 - Test |
| Eval2 | 4,264 | PubMed test split |
| Eval3 | 1,998 | LLM-generated questions, Gemini used |
| Total | 1,245,423 |  |

#### Data Sources and Preprocessing: Open Access PubMed Articles

We utilized the existing BIOMEDICA dataset [23], which contains image-caption pairs and metadata extracted from millions of open-access biomedical articles available on PubMed (PM). We filtered this dataset to identify figures relevant to dermatology using the keyword ‘Skin Lesion’ and selected samples with permissive licenses allowing research and derivative works (e.g., CC0, CC BY, CC BY-SA, CC BY-ND) (Supplementary Methods). From this filtered subset, we extracted 56,258 dermatology-related images after excluding non-skin images such as graphical figures. Each image was paired with its associated textual context, primarily figure captions and relevant inline mentions identified within the BIOMEDICA metadata, forming the basis for generating image-grounded question-answer pairs. We also developed a PubMed Central scrapping tool to collect up-to-date data of any field from the latest PubMed database (Supplementary Methods).

#### Data Collection and Preprocessing: Open-Access Datasets

We identified open-access datasets relevant to our target domain, dermatology, prioritizing collections with clear licensing terms and comprehensive documentation. For dermatology, this primarily included image repositories covering various skin conditions and demographic groups to enhance model generalizability. When necessary, images were resized or converted to consistent resolutions, and mislabelled or corrupted files were removed. Pertinent metadata—such as lesion type, imaging modality, or diagnostic tags—was extracted and centralized for easy reference. This coordinated pre-processing produced a unified, high-quality dataset suitable for both human annotation and automated synthetic data generation within the SCALEMED framework.

In this study, we employed DERM12345 [45], BCN20000 [46], HIBA [47], PAD-UFES-20 [48] and SCIN [8] datasets, all of which have commercial licenses suitable for use with SOTA models. These repositories collectively comprised 13,444 clinical images (30%) and 31,370 dermoscopic images (70%). Some datasets were accessed via the isic-cli library, while others required downloads from institutional or project-specific websites.

### Synthetic Data Generation

We employed multiple pipelines to generate a large and diverse synthetic dataset (DermaSynth) targeting various aspects of dermatological knowledge and reasoning. The generation process primarily utilized Google’s Gemini models (Gemini 2.0 Flash, from November 2024 to January 2025) via API calls. Gemini 2.0 Flash can generate high quality synthetic data even from basic seed tasks.

#### VQA Generation from Image Datasets (Open Access & Biomedica)

- **Open Access Datasets:** For each image in open-access datasets (e.g., DERM12345), we first converted their structured metadata into concise natural language strings (e.g., "Meta-data - Label: Melanoma, Age: 45, Sex: Female, Location: Forearm"). Predefined prompt templates included both "general" (e.g., "Describe the visual features in the image") and "dataset-specific" questions (e.g., "What is the diagnosis for this lesion located on the localization?") (Supplementary Methods). For each image, one general and one dataset-specific question were randomly selected. These questions and metadata strings were sent to Gemini. We requested two synthetic QA pairs per image, formatted as JSON format for easy parsing. From the open-access datasets, 92,020 samples were generated, with 2,392 excluded due to parsing issues.
- **BIOMEDICA Dataset:** For each selected BIOMEDICA sample, we combined its caption and associated contextual mentions into a single text block, which, along with the image, was provided to Gemini. Gemini rejected 14,429 images due to its guard system. The remaining 41,829 images were splitted into training and evaluation splits. We used two prompt styles: a "training style" prompt requesting three descriptive QA pairs regarding visible characteristics based on context, and an "evaluation style" prompt (used for generating Eval2) requesting either a multiple-choice or challenging open-ended question based on the image and context (Supplementary Methods).

#### Instruction Based Generation

We applied self-instruction techniques to generate diverse text-based and conversational data, beginning with small sets of human-curated seed tasks. Seed tasks were initially generated using ChatGPT o1 pro, then reviewed and edited by expert clinicians to ensure relevance and accuracy, including varied perspectives such as ‘explain like I am 5’. Our framework supports flexible integration of different seed tasks for diverse use cases, and we have designed it to be as comprehensive as possible.

1. Instruction-Following QA Synthesis: Starting with 190 seed task (Supplementary Data 1, 18 classification, 172 non-classification tasks, validated by clinicians Gulsum Gencoglan (G.G.), Ece Gokyayla (E.G.), and Ozan Erdem (O.E.)), we iteratively prompted Gemini (Supplementary Methods). In each iteration, two human-generated and two synthetic samples served as context. Gemini generated new instructions, labelling them as classification or non-classification tasks. For classification tasks, Gemini created class labels and inputs; for non-classification, it generated input-output pairs. Outputs were parsed using regular expressions, and near-duplicates were filtered using semantic similarity. 50,027 instruction-following samples were generated.
2. Knowledge-Base (KB) Task Synthesis: We developed a knowledge base covering approximately 2,700 dermatological diseases [49]. The knowledge base was sourced from Wikipedia’s list of skin conditions via Wikipedia-API (Supplementary Methods). We defined 40 seed task templates (e.g., "List key triggers for <DISEASE_NAME>", "Describe the typical presentation of <DISEASE_NAME>") (Supplementary Methods and Supplementary Data 2). For each disease, relevant templates and content sections were populated using disease information, extracted from the KB and used to prompt Gemini, which generated authoritative responses without directly referencing the provided context (Supplementary Methods). This approach generated 72,400 QA pairs grounded in dermatological knowledge.
3. Structured Case Synthesis (Anamnesis): We used 137 seed dermatological case studies based on the most common 137 dermatological disease (Supplementary Data 3-5), validated by clinicians and reflecting common conditions and doctor-patient interactions based on a standardized clinical questionnaire (17 sections, 104 questions) [29] (Supplementary Methods and Supplementary Data 3-5). By providing Gemini with example cases, we requested the generation of new case studies adhering to a strict JSON schema (sections such as base_information, questions_and_answers, final_diagnosis, and treatment_of_choice) (Supplementary Methods). Robust parsing handled variations in model outputs. This method generated 892,814 samples for 9,963 synthetic multi-turn case interactions.

#### Final DermaSynth Dataset

Outputs from these pipelines were aggregated to form the DermaSynth dataset, comprising approximately 1.2 million synthetic samples. All experiments were conducted to remain within a modest budget. The testing of GPT-4o was run on OpenAI API, yielding an inference cost of $100. Synthetic data creation via the Gemini API involved roughly 400,000 batched prompt-completion cycles. At the current rate of $0.55 per million output tokens, total generation cost was around $200. AnnotatorMed incurred no additional charges, hosted on a free Firebase plan. Thus, the entire pipeline, data generation, fine-tuning, and evaluation, was executed for well under $500, demonstrating that high-quality domain-specific models can be developed cost-effectively, accessible to most academic and clinical research groups.

### Evaluation Data, DermaBench

We created three distinct evaluation benchmarks to comprehensively assess model performance.

1. Eval1 (Expert-Curated Dermoscopic Image MCQA): Expert clinicians (G.G., E.G., O.E.) designed hierarchical questions (93 root and 320 total questions (Supplementary Data 6-8), including single-choice, multi-choice, and open-ended formats) based on the two-step pattern analysis widely used in skin lesion diagnosis (Supplementary Methods). They then annotated 225 images from the test split of the DERM12345 dataset (approximately five images per class across 40 classes) using these questions, creating a challenging VQA benchmark requiring nuanced clinical reasoning. In total, 93 main questions and 227 subquestions were developed, organized into five sections: intro, clinical, dermoscopy-basic, dermoscopy-melanocytic, dermoscopy-nonmelanocytic. The dermoscopic section follows the two-step pattern analysis [24]: Step 1 evaluates the basic lesion context. Step 2 classifies lesions as melanocytic or non-melanocytic, further subdivided into nine detailed levels. The summarization section similarly employs two steps: (1) evaluating global and local lesion patterns to classify as a benign nevus, suspect melanocytic lesion, or melanoma, and (2) determining the final diagnosis through specific decision questions. Questions irrelevant to dermoscopic images, such as certain clinical image inquiries, were omitted from Eval1 annotations. Additionally, each section includes two special questions prompting a summary report and requesting the model’s confidence level. To generate these annotations efficiently, we initially used the GPT-4o model to produce preliminary outputs, followed by multiple data validation steps to align the synthetic data with our backend and UI structures. These validation processes included removing invalid responses, correcting mislabelling (e.g., “Brown globules” → “globules”), verifying identifiers (ID), and ensuring all open-text fields, missing IDs, and option IDs adhered to the required format. A new thread was initiated in the OpenAI API for each image, with questions split appropriately to maintain them consistently. This method ensured synthetic annotations conformed to data structures while maintaining the high quality necessary for reliable evaluation.
2. Eval2 (Clinical Image MCQA): This benchmark comprises multiple-choice questions generated using an "evaluation style" prompt applied to the test split of the BIOMEDICA dataset. These questions assess comprehension and reasoning based on clinical images and their associated textual context, such as captions and inline mentions.
3. Eval3 (LLM-Generated Text-Only MCQA): Generated through a "Multiple-choice / balanced benchmark" pipeline, this benchmark consists of text-only multiple-choice questions covering various dermatology aspects, including diagnosis, treatment, and histopathology. Questions are balanced across difficulty levels (beginner, intermediate, expert), designed to assess the model’s textual dermatological knowledge base comprehensively.

### Model Training and Evaluation

In this section, we discuss the model training and evaluation strategies used to ensure robust performance across various use cases, supporting flexibility for both vision-based and text-based tasks. Depending on specific application requirements and the medical domain, either vision-based or text-based LLMs can be employed. Multi-modal models, which integrate vision, text, and other data types, offer further versatility for tasks requiring combined analysis of image and textual data. Text-based LLMs are particularly effective for structured data tasks, including medical record analysis, diagnostic reasoning, and clinical decision support. VLMs excel in image understanding and visual diagnostic applications. Choosing between these model types depends on data characteristics and specific medical domain needs, ensuring adaptability within diverse healthcare contexts. Different prompts were used for evaluating classification and report generation metrics. Additionally, single and multi-choice questions were used to calculate classification metrics, while all question types were used to calculate report generation metrics.

#### Teacher-Student Paradigm and Model Training

To balance performance and computational efficiency, we adopted a teacher-student approach. Initially, we utilized a larger teacher model, Gemini 2.0 Flash, to guide a lighter multimodal student model, Llama-3.2-11B-Vision in 16-bit precision (developed by Meta), distilling key insights through LoRA fine tuning [11]. This approach reduces hardware requirements while retaining high level reasoning capabilities. DermatoLlama models were trained and evaluated using Python (v3.10.13), LLaMa-Factory (v0.9.2) [50] and DeepSpeed (v0.16.3). Training was performed on a low-cost workstation equipped with four Nvidia RTX 4090 GPUs (96 GB combined VRAM) and 512 GB system RAM. For inference and benchmarking, we used a computer with a single RTX 4090 GPU (24 GB VRAM) and 96 GB RAM, confirming practical deployment feasibility on a single-GPU setup.

#### Low-Rank Adaptation (LoRA) Fine-Tuning

Pretraining or full finetuning of LLMs involves updating all weights, a memory-intensive and computationally demanding process. In contrast, LoRA fine-tuning [11] updates only a small, low-rank matrices.

For any linear layer with pretrained weights **W**_0_, LoRA adds a learnable correction

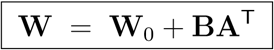

where **A** ∈ ℝ*^k×r^* and **B** ∈ ℝ*^d×r^* have a very small *rank r*.

In practice, we applied LoRA to DermatoLlama with a rank of 16, dropout rate of 0.05, and input/output cut-off of 2,048 for targeted modules (Supplementary Methods). During fine-tuning, only the LoRA matrices (A and B) were updated. After training, these low-rank updates were merged back into the original weight matrices, ensuring no additional memory or latency overhead upon deployment. This strategy achieved SOTA dermatology classification while fitting comfortably on a single RTX 4090 GPU for inference.

After training, the low-rank products were merged into **W**_0_ for zero-overhead inference, or maintained separately, allowing a single base model to support multiple domain-specific LoRA adapters with minimal storage requirements.

#### Model Auditing

To verify model improvements, we incrementally introduced samples from our synthetic dataset, DermaSynth, at scales of 10k, 50k, 100k, 200k, and Full dataset, assessing performance at each increment. The test split of DERM12345 was excluded from training. Model performances at each stage were evaluated using the evaluation datasets (Eval1, Eval2, Eval3), measuring accuracy, BLEU-4, ROUGE-1, ROUGE-2, and ROUGE-L scores to assess DermatoLlama variants and SOTA models.

### Tools

We developed AnnotatorMed, an open-source, lightweight annotation platform built using Flutter (v3.22.3), and deployable on both web and mobile devices. A Firebase backend manages labelling records; however, any database can be integrated for on-premises installations. Additionally, we developed a PubMed scrapping tool designed to filter specific fields from the PubMed Central database (Supplementary Methods).

AnnotatorMed features a graphical interface structured around a hierarchical data model-question → option → sub-question → sub-option-that simplifies nested clinical logic into an intuitive single screen flow. This approach supports three question formats: single-choice, multi-choice, and open-ended. Selecting an option reveals only relevant follow-up items, significantly reducing user interactions and cognitive load. The same component library can also operate in a flat, non-hierarchical mode if preferred. All annotations can be exported as CSV or JSON, facilitating seamless integration into downstream fine-tuning and evaluation pipelines. Using AnnotatorMed, three dermatologists (G.G., E.G., O.E.) labelled 225 images from the DERM12345 test split. The intuitive, expand-on-demand interface reduced annotation time while ensuring all decisions-including clinical findings, dermoscopic cues, and free-text comments-were stored consistently in machine-readable format.

### License Issues

All synthetic data generated from PubMed articles and open-access datasets used in this study allow for commercial usage. However, data generation was utilized using OpenAI and Gemini APIs, each subject to their own usage restrictions. The VLM model, DermatoLllama, was fine-tuned using Llama-3.2-11B-Vision model. DermaSynth and DermatoLlama are released under CC-BY-NC 4.0 license, prohibiting direct commercial use. This aligns with:

- Llama’s Non-Commercial License: Derivative projects inherit non-commercial restrictions.
- Upstream Model Limitations: Some instructional data originate from providers that prohibit commercial use.
- Safety and Maturity Concerns: Additional safeguards are necessary before broader clinical deployment.

Researchers must independently verify compliance with all upstream licenses and implement appropriate safety measures before adopting these models in real-world patient-care scenarios.

## Data Availability

BIOMEDICA dataset is publicly available from https://huggingface.co/BIOMEDICA. Open access datasets are all publicly available and can be accessed from: DERM12345 (https://dataverse.harvard.edu/dataset.xhtml?persistentId=doi:10.7910/DVN/DAXZ7P), BCN20000 (https://figshare.com/articles/journal_contribution/BCN20000_Dermoscopic_Lesions_in_ the_Wild/24140028/1), SCIN (https://github.com/google-research-datasets/scin?tab=readme-ov-file), HIBA (https://api.isic-archive.com/collections/175/), and PAD-UFES-20 (https://data.mendeley.com/datasets/zr7vgbcyr2/1). DermaSynth dataset with DermaBench and DermatoLlama models are available at https://huggingface.co/DermaVLM.

## Code Availability

Codes are available at https://github.com/DermaVLM.

## Acknowledgement

Abdurrahim Yilmaz has been funded by the President’s PhD Scholarships at Imperial College London. Donghee Choi and Joram M. Posma are supported by the Horizon Europe project CoDiet. The CoDiet project is funded by the European Union under Horizon Europe grant number 101084642. CoDiet research activities taking place at Imperial College London is supported by UK Research and Innovation (UKRI) under the UK government’s Horizon Europe funding guarantee (grant number 101084642). All figures were created with BioRender.

